# Effectiveness of SGLT2 inhibitor therapy in treatment of Heart failure: A Meta-Analysis

**DOI:** 10.1101/2023.09.10.23295313

**Authors:** Sarvinoz Albalushi, Azmaeen Zarif, Suheyla Karaduman, Khoa Tran, Alesia Talpeka

**Affiliations:** University of Debrecan, Debrecen, Egyetem tér 1, 4032 Hungary; University of Cambridge School of Clinical Medicine, Addenbrooke’s Hospital, Hills Road, Cambridge, Cambridgeshire, CB2 0SP, United Kingdom; Gonville & Caius College, University of Cambridge, Trinity Street, Cambridge, Cambridgeshire, CB2 1TA, United Kingdom; Uludag University Medical School, Özlüce, Uludağ Ünv., 16285 Nilüfer/Bursa, Türkiye; Pham Ngoc Thach University of Medicine, 2 Ð. Dương Quang Trung, Phường 12, Quận 10, Thành phố Hố Chí Minh, Vietnam; Belarusian State Medical University, Prospekt Dzerzhinskogo 83, Minsk 220116, Belarus; Minsk City Clinical Oncology Center, 1st City Clinical Hospital, Nezavisimosti Svenue, 64, Minsk 220040

**Author notes:** Corresponding author: Azmaeen Zarif, Gonville & Caius College, University of Cambridge, Trinity Street, Cambridge, Cambridgeshire, CB2 1TA, United Kingdom. Both SA and AZ contributed equally to this work as co-first-authors. SK, KT, and AT undertook the literature search and helped prepare the manuscript draft. All authors read and approved the final manuscript.

**Keywords:** sodium-glucose co-transporter 2 inhibitors, heart failure with preserved ejection fraction, heart failure with reduced ejection fraction, hospitalization, mortality

## Abstract

**Aim:** We conducted a meta-analysis on the effectiveness of sodium-glucose co-transporter 2 (SGLT2) inhibitors in heart failure (HF) patients with preserved (HFpEF) and reduced (HFrEF) ejection fraction.

**Methods:** A comprehensive search of several databases for placebo-controlled randomised controlled trials of SGLT2 inhibitors from inception to September 30 2022, limited to English language and excluding animal studies, was conducted. The databases included Medline, Scopus, Cochrane Central, and ClinicalTrials.gov. Randomised controlled studies were chosen if they included HF patients and reported at least one of the predetermined outcomes. Hazard ratios (HRs) or risk ratios were pooled together with the appropriate 95% confidence intervals using a random-effect model. Fourteen trials met the inclusion criteria, with 42 409 HF patients participating (N = 21 678 in the SGLT2 inhibitor arms and N = 20 731 in the placebo arms).

**Results:** SGLT2 inhibitors significantly reduced the composite of HF hospitalisation or cardiovascular death [HR: 0.75 (0.71–0.79); *P* < 0.001; *I*^2^ = 0%], total hospitalisations for heart failure [HR: 0.70 (0.66– 0.75); *P* < 0.001; *I*^2^ = 0%], and reduced the occurrence of cardiovascular death [HR: 0.86 (0.81–0.92); *P* < 0.001; *I*^2^ = 43%]. Sub-group analysis was conducted based on HF status at baseline, EF, diabetes status at baseline, and sex. SGLT2 inhibitors significantly reduced the composite heart failure hospitalisation and cardiovascular death for all the subgroups except for patients with HFpEF at baseline [HR: 0.87 (0.74– 1.03); *P* = 0.11; *I*^2^ = 60%] and no diabetes status at baseline [HR: 0.92 (0.73–1.16); *P* = 0.50; *I*^2^ = 78%].

**Conclusions:** In patients with HF, SGLT2 inhibitors may significantly improve clinical outcomes, including all-cause and cardiovascular mortality. In this meta-analysis, SGLT2 inhibitors did not show a reduction in composite heart failure in patients with heart failure with preserved ejection fraction at baseline.

## Introduction

Sodium-glucose co-transporter inhibitor (SGLT2i) has emerged to be effective in the treatment of heart failure with preserved (HFpEF) and reduced ejection fraction (HFrEF). Recently, The American Heart Association (AHA), The American College of Cardiology (ACC), and Heart Failure Society of America (HSFA) published guidelines for the management of heart failure in which SGLT2i was introduced as a novel therapeutic drug class (1,2). Careful evaluation of new evidence and comprehensive literature research encompassing clinical trials, research studies, and review papers revealed its efficacy and benefit in treating HFrEF to reduce hospitalisation. Since an earlier meta-analysis (3), covering seven major trials, was published in 2020, there have been seven more major trials that have been conducted (4-10). Therefore, we conducted this meta-analysis to provide a revised estimation of the efficacy of SGLT2i in heart failure patients.

## Method

### Data sources and search strategy

Cochrane and PRISMA (Preferred Reporting Items for Systematic review and Meta-Analyses) guidelines were utilised (11,12). A comprehensive search of several databases for placebo-controlled randomised controlled trials of SGLT2 inhibitors from inception to September 30 2022, limited to English language and excluding animal studies, was conducted. The databases included: Medline, Scopus, Cochrane Central, and ClinicalTrials.gov. In order to find relevant results, the following keywords and their MeSH terms were used: (Sodium Glucose Transporter 2 Inhibitors OR sodium-glucose co-transporter inhibitor OR SGLT-2 Inhibitors OR SGLT 2 Inhibitors OR SGLT2 Inhibitors OR Gliflozins OR tofogliflozin OR sotagliflozin OR empagliflozin OR canagliflozin OR dapagliflozin OR ertugliflozin OR luseugliflozin OR ipragliflozin OR remogliflozin OR sergliflozin AND Cardiac Failure OR Myocardial Failure OR Congestive Heart Failure OR heart failure OR Heart Decompensation). See Table S1 for the specific search strategies utilised for the respective databases. For the purpose of finding any pertinent studies that may have been missed during the search, reference lists of the included trials were carefully checked.

### Selection criteria for the study and eligibility

Articles were chosen for inclusion if they complied with all of the following eligibility requirements: HF patients were included, the studies (i) compared SGLT2 inhibitors to placebo, (ii) randomised control trial (RCT) or post hoc assessments of RCTs, and (iii) contained at least one of the specified outcomes of interest.

### Data extraction, outcomes of interest, and quality evaluation

The composite of cardiovascular death or HF hospitalisation was the major outcome of interest. Other results of interest were: (i) composite of total HF hospitalisation and cardiovascular death; (ii) cardiovascular death; (iii) death with HF at baseline; (iv) all-cause death; (v) renal composite outcome among all patients; and (vi) renal composite outcome based on HF status at baseline. A sub-group analysis based on baseline HF status, ejection fraction, baseline diabetes status, and baseline sex was also conducted.

### Statistical analysis

All statistical analyses were performed using RevMan (Review Manager Version 5.4), Copenhagen: The Nordic Cochrane Centre, (The Cochrane Collaboration, 2014). For four time-to-event clinical outcomes, including Composite of total HF hospitalisation or cardiovascular death, hospitalisation for heart failure, cardiovascular death, all-cause mortality, and composite renal outcome, hazard ratios and accompanying 95% confidence intervals were retrieved. Hazard ratios were log transformed and pooled using either fixed effect or random effect model. Using the DerSimonian-Liard random-effect model, we accounted for the expected heterogeneity in the study design, difference in the intervention drug and its usage, and variance in operationalisation of outcome. Where heterogeneity was less than 50%, a fixed-effect model was used instead. We applied an inverse variance method to assign the respective weighting of included studies. In a random-effect model, the weight of a study is equal to the inverse of the variance of the study effect and allows the study outcomes to vary according to a normal distribution. However, in a fixed-effect model, the weight given to each study in the analysis varies according to the sample size.

A subgroup analysis was conducted based on sex, diabetes status at baseline, heart failure at baseline (this cohort included all patients with HFrEF, HFpEF, and patients with unknown type of heart failure), and for patients with HFrEF, and HFpEF.

Subgroup differences were analysed using *χ*^2^ test while statistical heterogeneity was measured via *I*^2^ statistical testing. We used post-hoc analysis statistics of trials for subgroup analysis. Publication bias was analysed using funnel plot. Statistical significance was defined as a P-value ≤0.05.

## Result

### Study Characteristics

The procedure for searching and choosing studies is summarised in the PRISMA flow chart (Supplementary Materials, Figure S1). Data were gathered from 14 RCTs that assessed SGLT2 inhibitors. 42 409 HF patients participated in the total trial population for this meta-analysis (N = 21 678 in the SGLT2 inhibitor arms and N = 20 731 in the placebo arms). Of those, 18 838 participants made up the HFrEF subgroup (N = 9475 in the SGLT2 arms and N = 9363 in the placebo arms). There were 8542 participants in the HFpEF subgroup (N = 4441 in the SGLT2 arm and N = 4101 in the placebo arm). The HF type for the remaining individuals were not indicated in their respective studies.

**Table 1.**
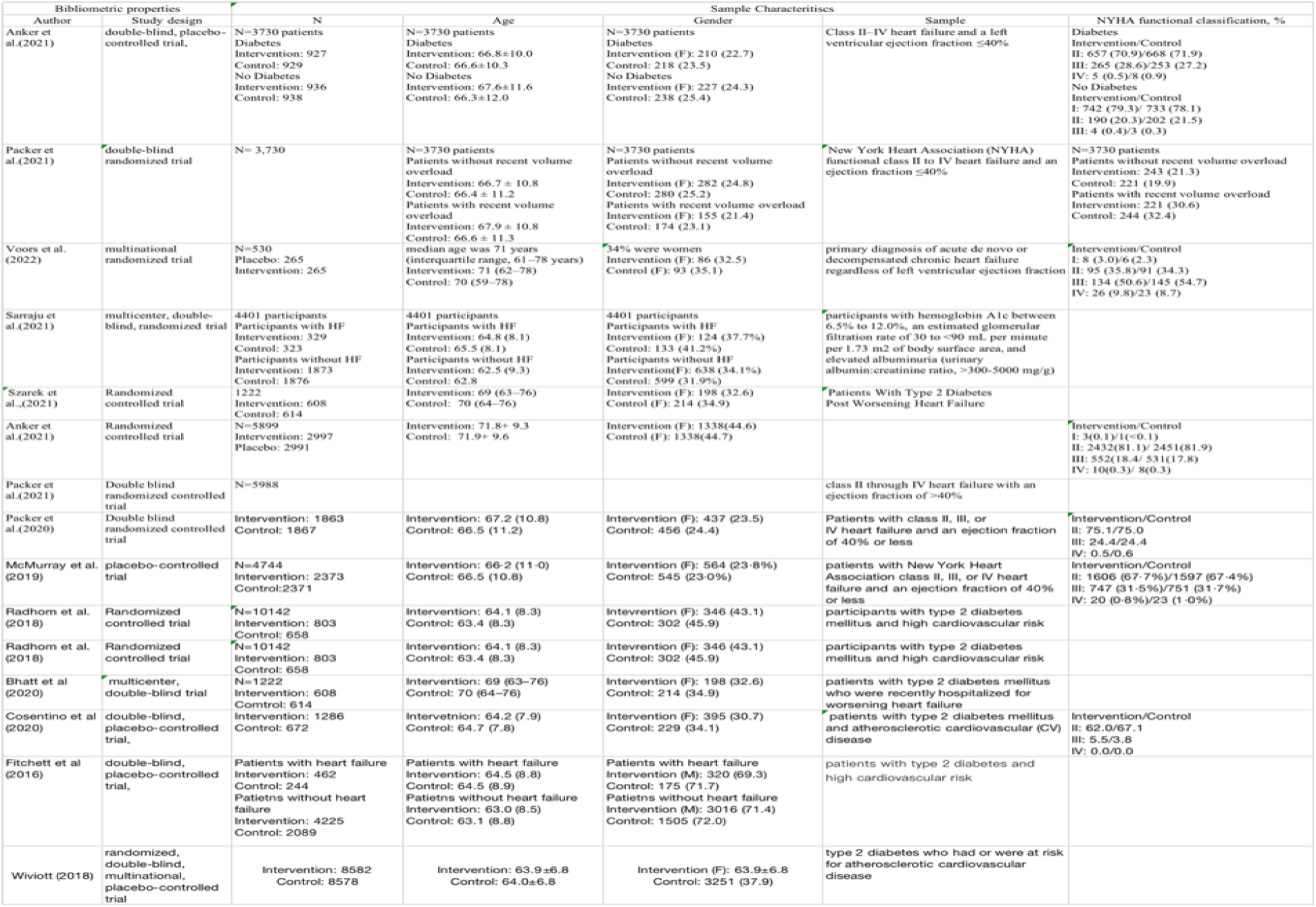
Characteristics of selected studies.

**Table 2.**
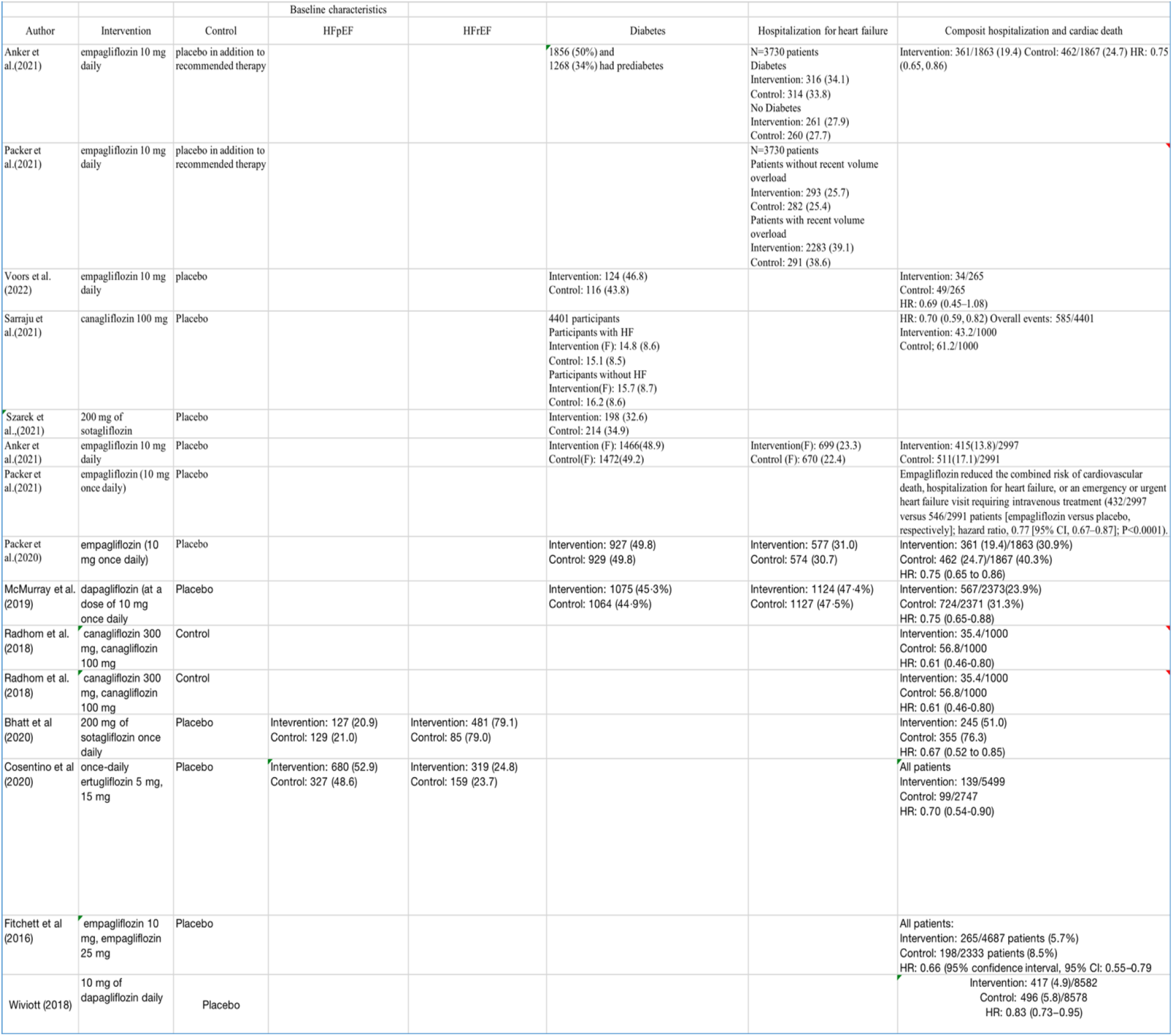
Baseline patient characteristics in selected studies.

**Table 3.** Summary clinical outcomes reported from selected studies.

| Author | Outcome |  |  |  |
| --- | --- | --- | --- | --- |
|  | Hospitalization for heart failure | Cardiac death | All cause death | Renal composite |
| Anker et al.(2021) | Intervention: 388/1863 Control: 553/1867 HR: 0.70 (0.58, 0.85) | Intervention: 187/1863 (10.0) Control: 202/1867 (10.8) HR: 0.92 (0.75, 1.12) |  | Intervention: 30/1863 (1.6) Control: 58/1867 (3.1) HR: 0.50 (0.32, 0.77) HR: 0.50 (0.32, 0.77) |
| Packer et al.(2021) |  |  |  |  |
| Voors et al. (2022) | Intervention (14 (5.3)/265)<br>Control (12 (4.5)/265)<br>HR: 1.179 (0.534–2.601) | Intervention: 11 (4.2)/265<br>Control: 22 (8.3)/265 |  |  |
| Saraju et al.(2021) | Overall events: 230/4401<br>Intervention: 15.7/1000<br>Control: 25.3/1000<br>HR: 0.61 (0.47, 0.80) | Overall events: 486/4401<br>Intervention: 38.7/1000<br>Control: 48.7/1000<br>HR: 0.80 (0.67, 0.95) | Overall events: 369/4401<br>Intervention: 29.0/1000<br>Control: 35.0/1000<br>HR: 0.83 (0.68, 1.02) | Overall events: 377/4401<br>Intervention: 27.0/1000<br>Control: 40.0/1000<br>HR: 0.66 (0.53, 0.81) |
| Szarek et al.,(2021) | HR: 0.76 (0.63–0.93)<br>Intervention: 97.6 (83.4–111.8)/608<br>Control: 126.6 (107.6–145.6)/614 |  |  |  |
| Anker et al.(2021) | Intervention:259(8.6)/ 2997<br>control: 352(11.8)/ 2991 | intervention : 219(7.3)/ 2997<br>Control: 244(8.2)/ 2991 | Intervention: 422(14.1)/ 2997<br>Control: 427(14.3)/2991 | Intervention: 108(3.6)/ 2997<br>control: 112(3.7)/ 2991 |
| Packer et al.(2021) | Empagliflozin reduced the total number of heart failure hospitalizations that required intensive care (hazard ratio, 0.71 [95% CI, 0.52–0.96]; P=0.028) [(86/2997 versus 117/2991 patients [empagliflozin versus placebo, respectively] |  |  | Compared with patients in the placebo group, fewer patients in the empagliflozin group reported outpatient intensification of diuretics (482/2997 versus 610/2991; hazard ratio, 0.76 [95% CI, 0.67–0.86]; P<0.0001), |
| Packer et al.(2020) | Intervention:388/1863<br>Control: 553/1867<br>HR: 0.70 (0.58 to 0.85) | Intervention: 187/1863 (10.0%)<br>Control: 202/1867 (10.8%)<br>HR: 0.92 (0.77-1.12) | Intervention: 249/1863 (13.4%)<br>Control: 266/1867 (14.2%)<br>HR: 0.92 (0.77-1.10) | Intervention:30 (1.6)/1863<br>Control: 58 (3.1)/1867<br>HR: 0.50 (0.32 to 0.77) |
| McMurray et al. (2019) | Intervention: 231/2373(9.7%)<br>Control: 318/2371 (13.4%)<br>HR: 0.70 (0.59-0.83) | Intervention: 227/2373(9.6%)<br>Control: 273/2371 (11.5%)<br>HR: 0.82 (0.69-0.98) | Intervention: 276/2373 (11.6%)<br>Control: 329/2371 (13.9%)<br>HR: 0.83 (0.71-0.97) | Intervention: 28/2373(1.2%)<br>Control: 39/2371 (1.6%)<br>HR: 0.71 (0.44-1.16) |
| Radhom et al. (2018) | Intervention: 14.1/1000<br>Control: 28.1/1000<br>HR: 0.51 (0.33-0.78) | Intervention: 24.3/1000<br>Control: 31.6/1000<br>HR: 0.72 (0.51-1.02) | Intervention: 29.2/1000<br>Control: 38.7/1000<br>HR: 0.70 (0.51-0.96) | Intervention: 6.8/1000<br>Control: 11.0/1000<br>HR: 0.67 (0.30-1.51) |
| Radhom et al. (2018) | Intervention: 14.1/1000<br>Control: 28.1/1000<br>HR: 0.51 (0.33-0.78) | Intervention: 24.3/1000<br>Control: 31.6/1000<br>HR: 0.72 (0.51-1.02) | Intervention: 29.2/1000<br>Control: 38.7/1000<br>HR: 0.70 (0.51-0.96) | Intervention: 6.8/1000<br>Control: 11.0/1000<br>HR: 0.67 (0.30-1.51) |
| Bhatt et al (2020) | Intervention: 194 (40.4)<br>Control: 297 (63.9)<br>HR: 0.64 (0.49 to 0.83) | Intervention: 51 (10.6)<br>Control: 58 (12.5)<br>HR: 0.84 (0.58 to 1.22) | Intervention: 65 (13.5)<br>Control: 76 (16.3)<br>HR: 0.82 (0.59 to 1.14) |  |
| Cosentino et al (2020) |  |  |  |  |
| Fitchett et al (2016) | All patients:<br>Intervention: 126/4687 patients (2.7%)<br>Control: 95/2333 patients (4.1%)<br>HR: 0.65 (95% CI: 0.50–0.85) | All patients:<br>Intervention: 172/4687 patients<br>Control: 137/2333 patients<br>HR: 0.62 (95% CI: 0.49–0.77) | All patients:<br>Intervention: 269/4687 patients<br>Control: 194/2333 patients<br>HR: 0.68 (95% CI: 0.57–0.82) |  |
| Wiviott (2018) | Intervention: 212 (2.5)/8582<br>Control: 286 (3.3)/8578<br>HR: 0.73 (0.61–0.88) | Intervention: 245 (2.9)/8582<br>Control: 249 (2.9)/8578<br>HR: 0.93 (0.82–1.04) | Intervention: 529 (6.2)/8582<br>Control: 570 (6.6)/8578<br>HR: 0.98 (0.82–1.17) |  |

**Table 4.**
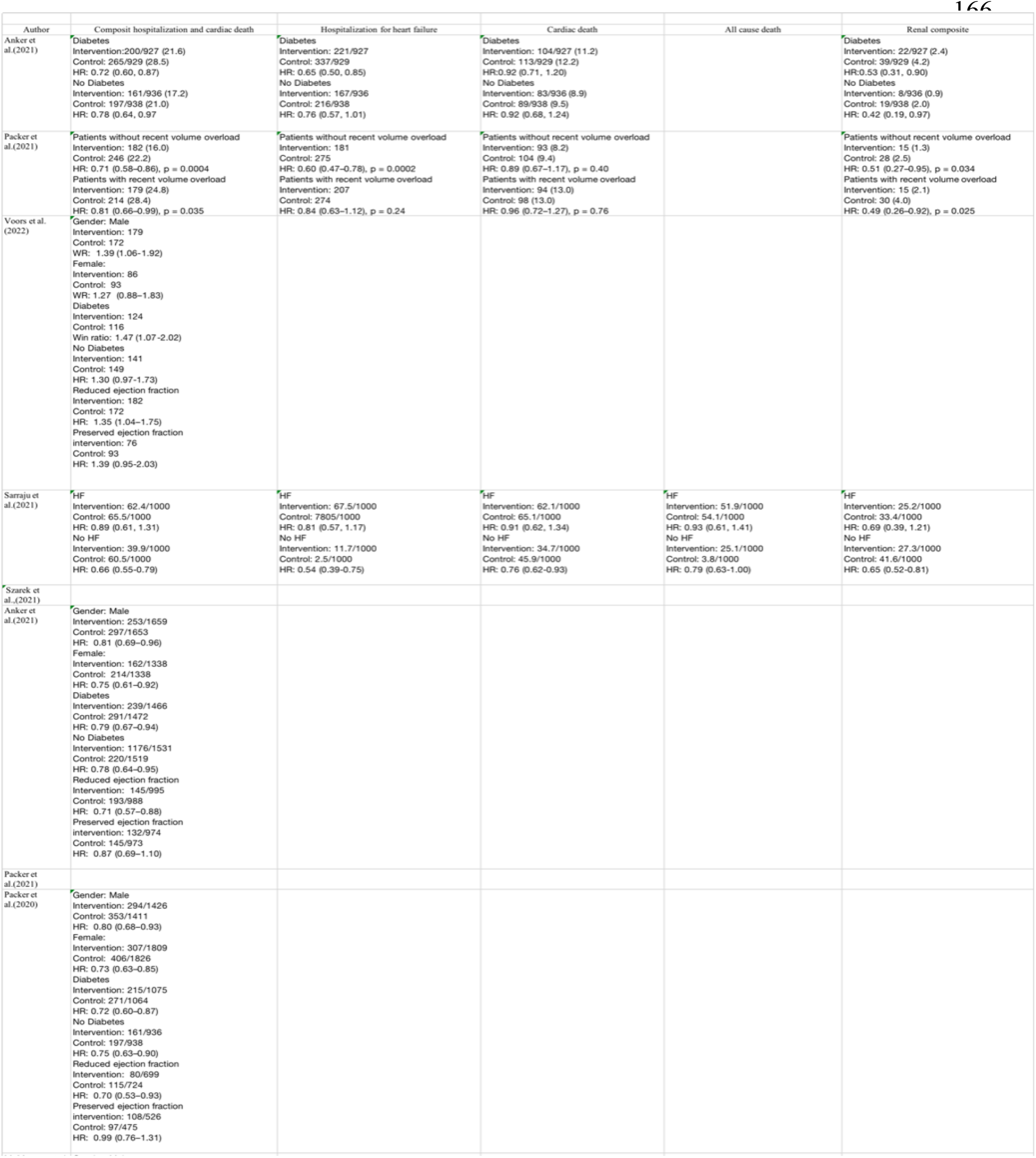
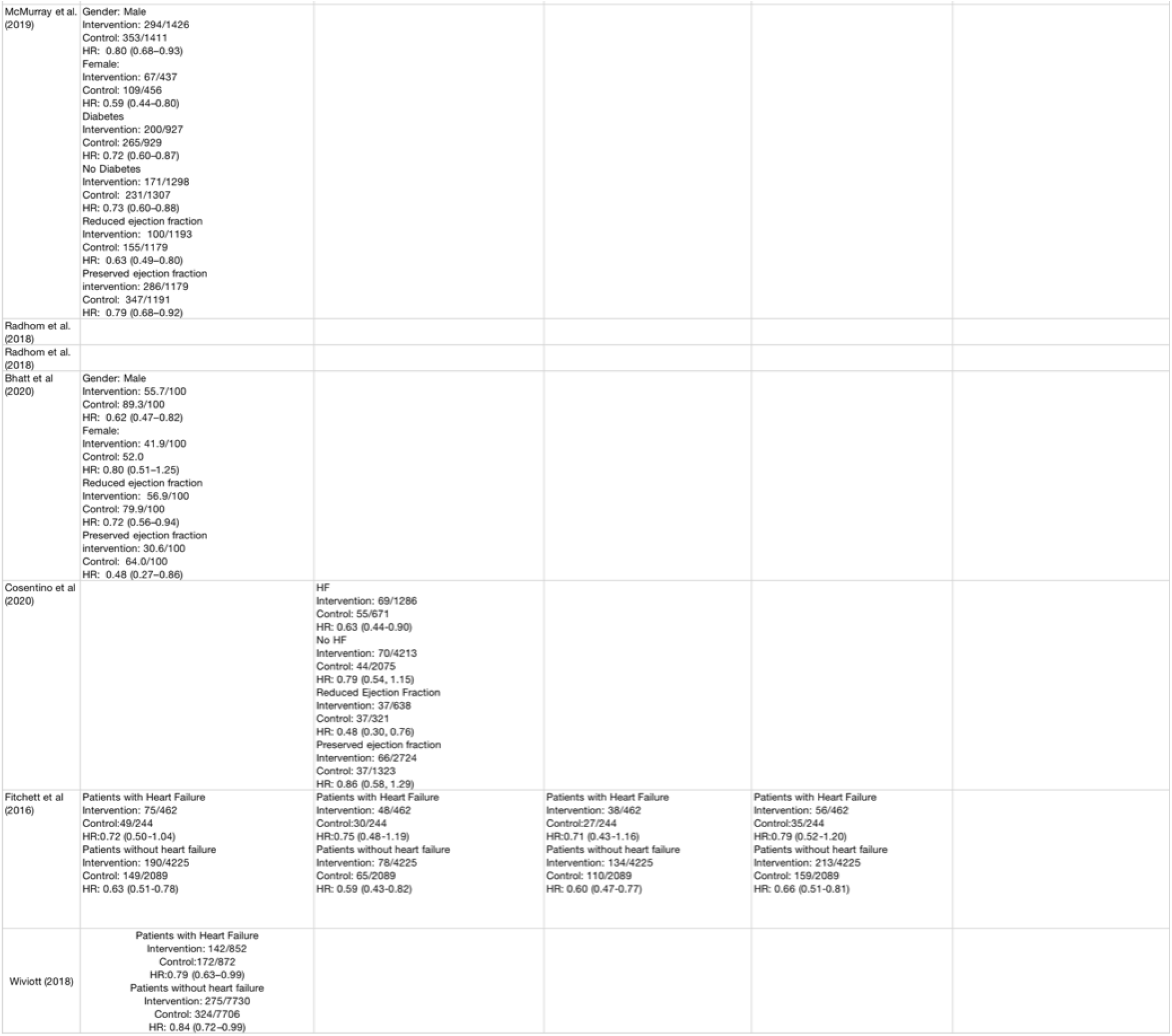
Sub-group analysis clinical outcomes. ARB: angiotensin receptor blocker; ARNI: angiotensin receptor/neprilysin inhibitor; ACE: angiotensin-converting enzyme; CRT-P: cardiac resynchronisation therapy pacemaker; eGFR: estimated glomerular filtration rate; ICD: implantable cardiac defibrillator; LVEF: left ventricular ejection fraction; NT-proBNP: N-terminal pro-B type natriuretic peptide; NYHA: New York Heart Association; SD: standard deviation; HFpEF: heart failure with preserved ejection fraction; HFrEF: heart failure with reduced ejection fraction. Information is presented as n (%), mean (SD), or median (interquartile range).

## Results

### Composite of HF hospitalization or cardiac death

Results of meta-analysis indicated that SGLT2 inhibitors significantly reduced the composite of HF hospitalisation or cardiovascular death [HR: 0.75 (0.71–0.79); P < 0.001; I^2^ = 0%] (Figure 1). Sub-group analysis was conducted to based on HF status at baseline, ejection fraction, diabetes status at baseline, and gender. SGLT2 inhibitors significantly reduced the composite heart failure hospitalisation and cardiac death for all the subgroups except for patients with HFpEF at baseline [HR: 0.87 (0.74–1.03); P = 0.11; I^2^ = 60%] and no diabetes status at baseline [HR: 0.92 (0.73–1.16); P = 0.50; I^2^ = 78%]. Moreover, there was significant difference between patients with and without heart failure at baseline [*χ*2 = 0.06, df = 1; P = 0.81), I^2^= 0%], patients with reduced and preserved ejection fraction [*χ*2 = 0.08, df = 1; P = 0.37), I2= 0%], patients with and without diabetes status at baseline [*χ*2 = 0.62, df = 1; P = 0.43), I^2^= 0%], and men and women [*χ*2 = 0.34, df = 1; P = 0.56), I^2^= 0%]. Sub-group analyses are presented in Figure 2.

**Figure 1.**
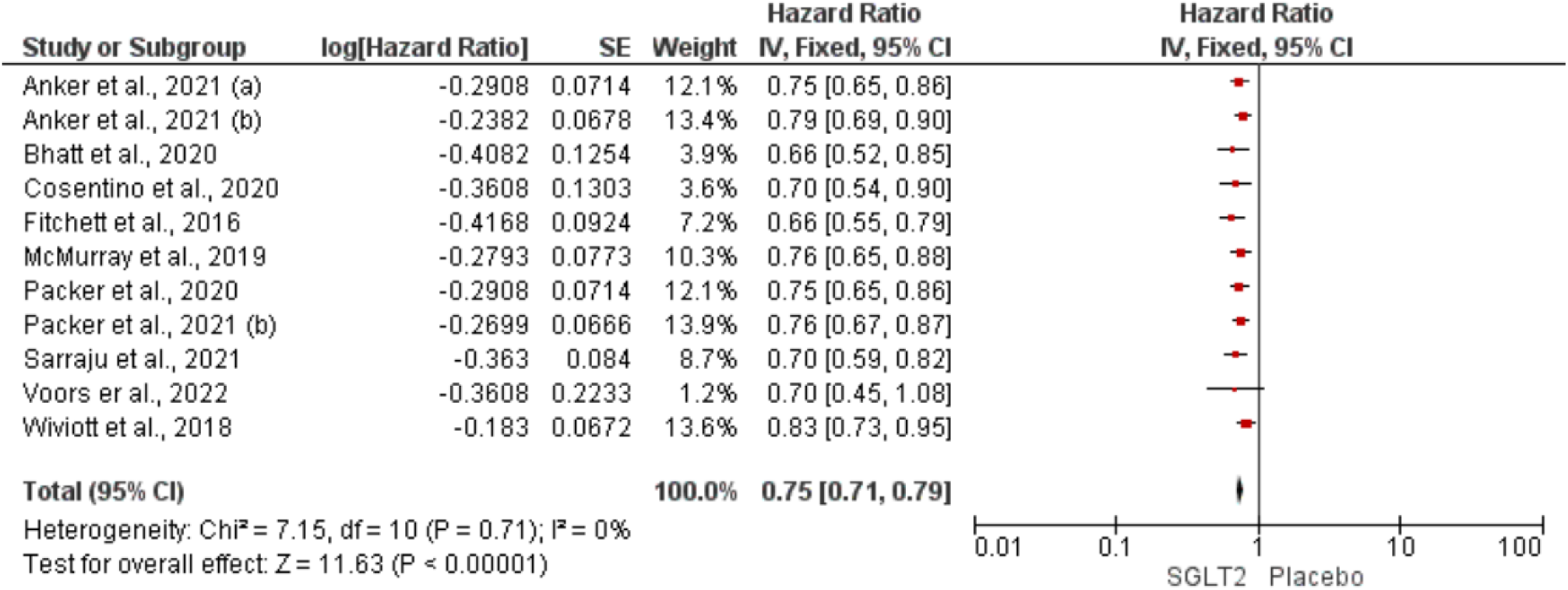
Forest plot displaying effect size for composite score of HF hospitalisation or cardiac death among all patients

**Figure 2.**
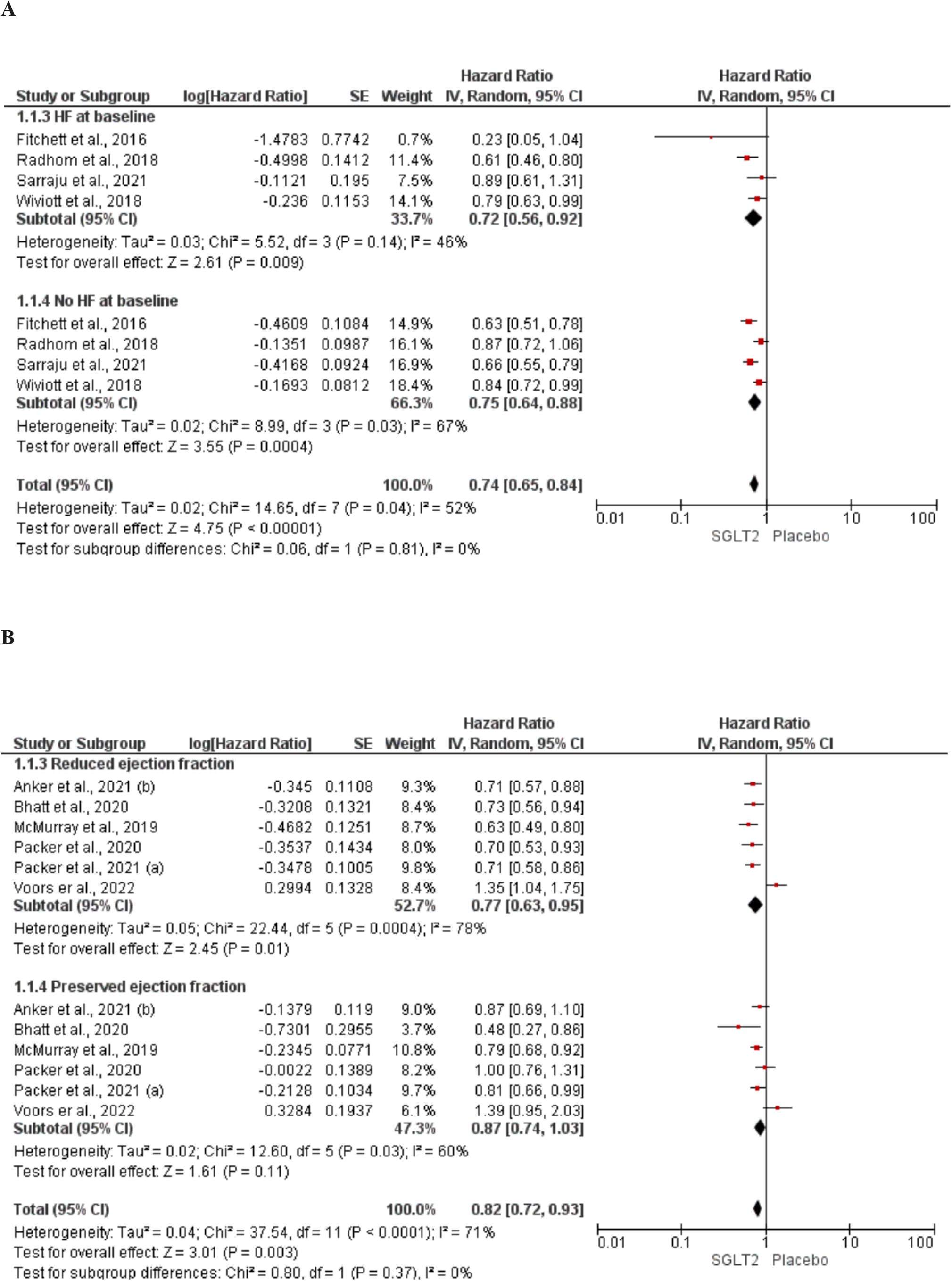
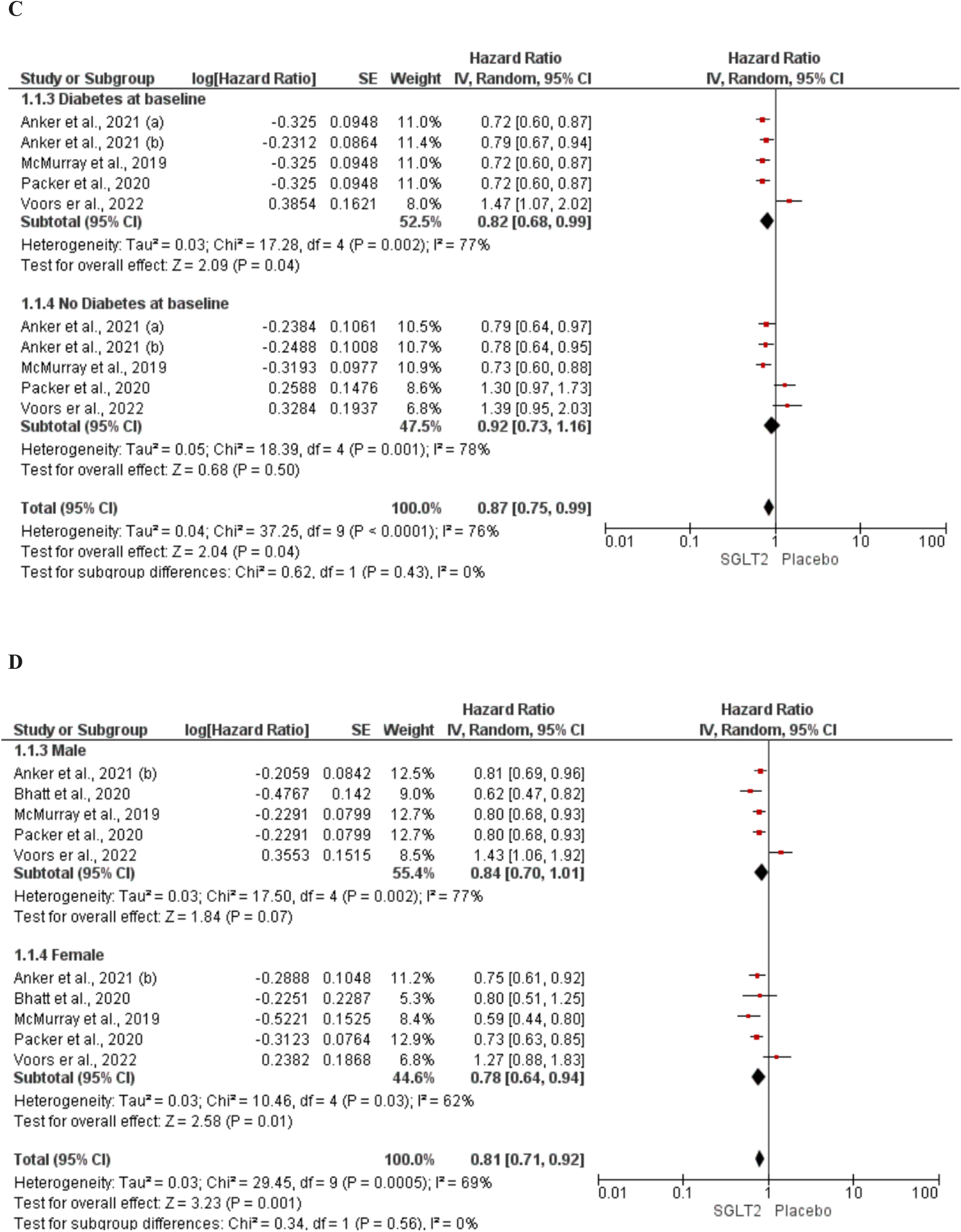
Sub-group analysis for composite score of HF hospitalisation or cardiac death based on HF status at baseline (**A**), HFrEF & HFpEF (**B**), diabetes status at baseline (**C**), and gender (**D**).

### Hospitalization for heart failure (HHF)

SGLT2 inhibitors resulted in a significant reduction in total hospitalisations for heart failure [HR: 0.70 (0.66– 0.75); P < 0.001; I^2^ = 0%] (Figure 3). Sub-group analysis was conducted for patients with and without heart failure at baseline and results indicated that SGLT2 inhibitors reduced hospitalisation for heart failure among both group with no significant difference between them [*χ*2 = 0.02, df = 1; P = 0.89), I^2^= 0%] (Figure 4).

**Figure 3.**
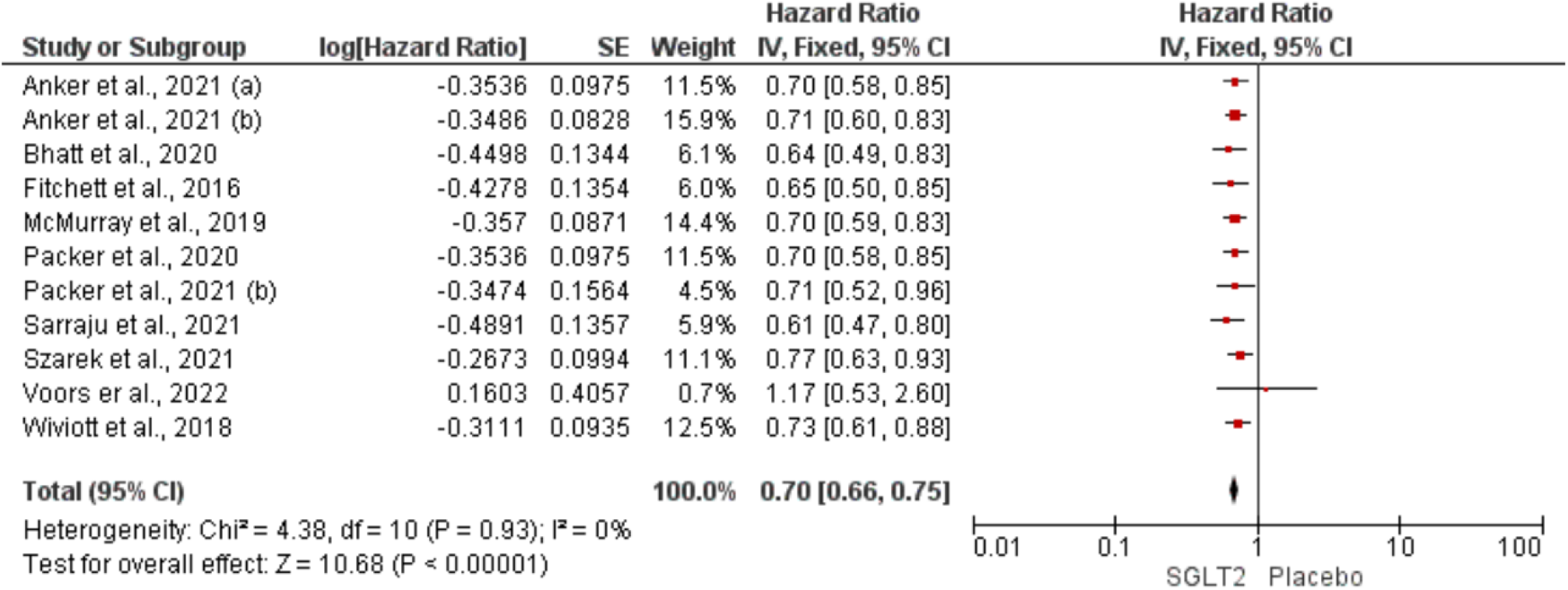
Forest plot displaying effect size for HHF.

**Figure 4.**
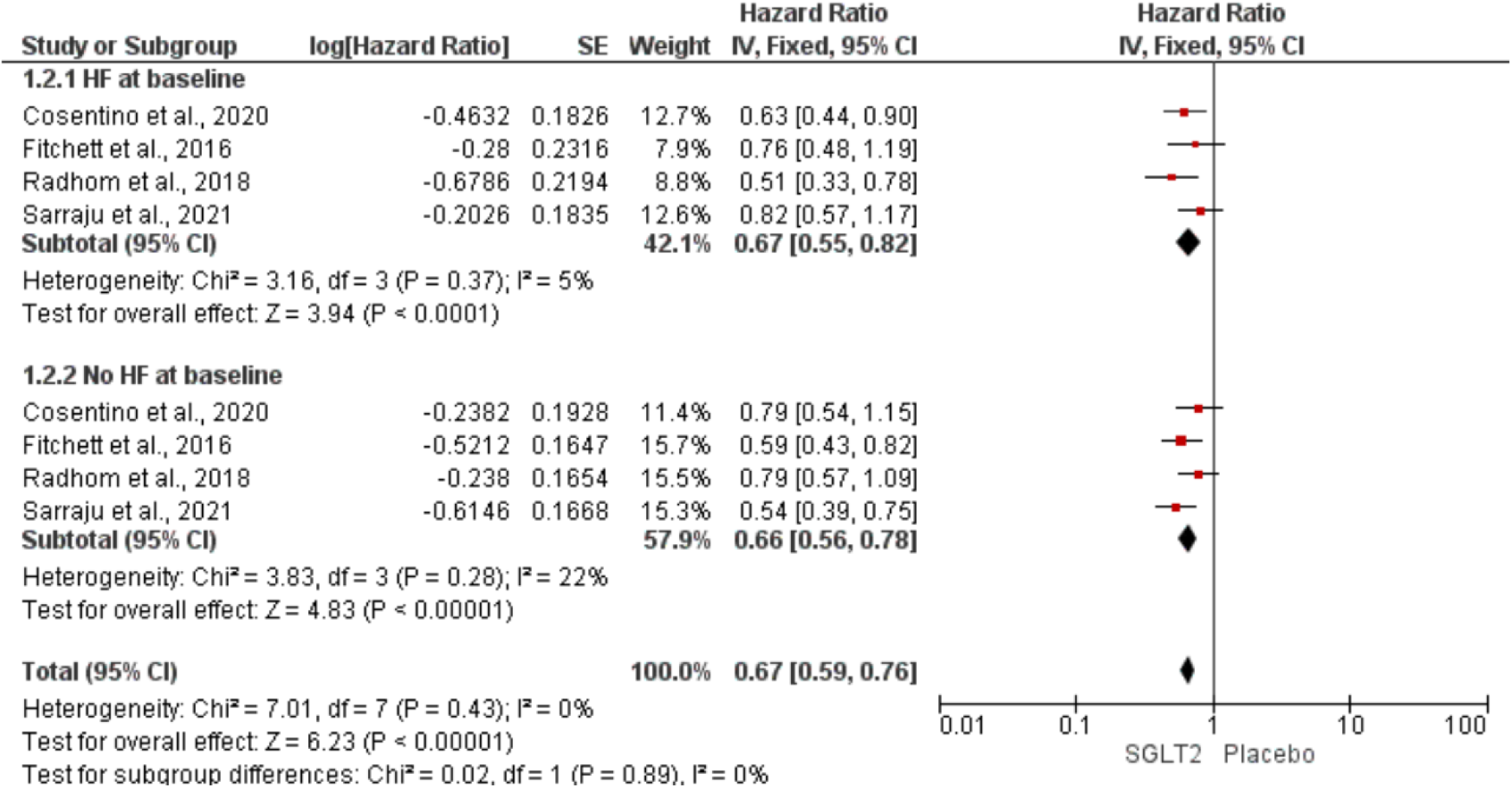
Sub-group analysis for HHF based on HF status at baseline.

### Cardiac Death

SGLT2 inhibitors significantly reduced the occurrence of cardiovascular death among participants [HR: 0.86 (0.81– 0.92); P < 0.001; I^2^ = 43%] (Figure 5). The findings were consistent in patients with and without heart failure at baseline [*χ*2 = 0.01, df = 1; P = 0.97), I^2^= 0%] depicted in sub-group analysis Figure 6.

**Figure 5.**
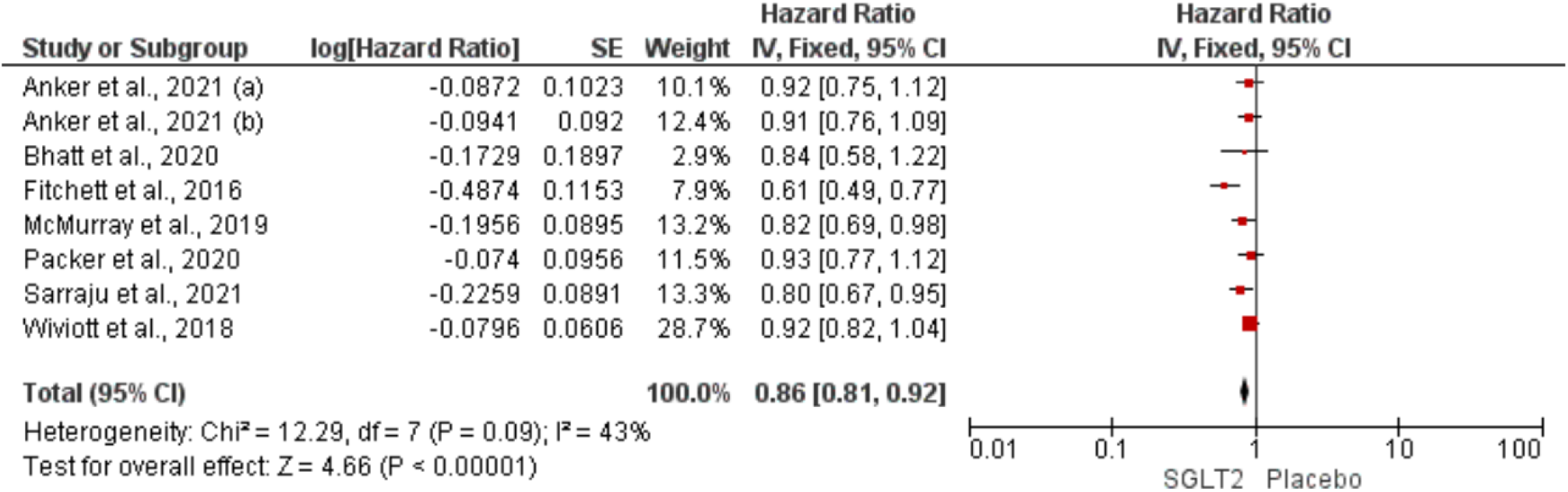
Forest plot displaying effect size for cardiac death among all patients.

**Figure 6.**
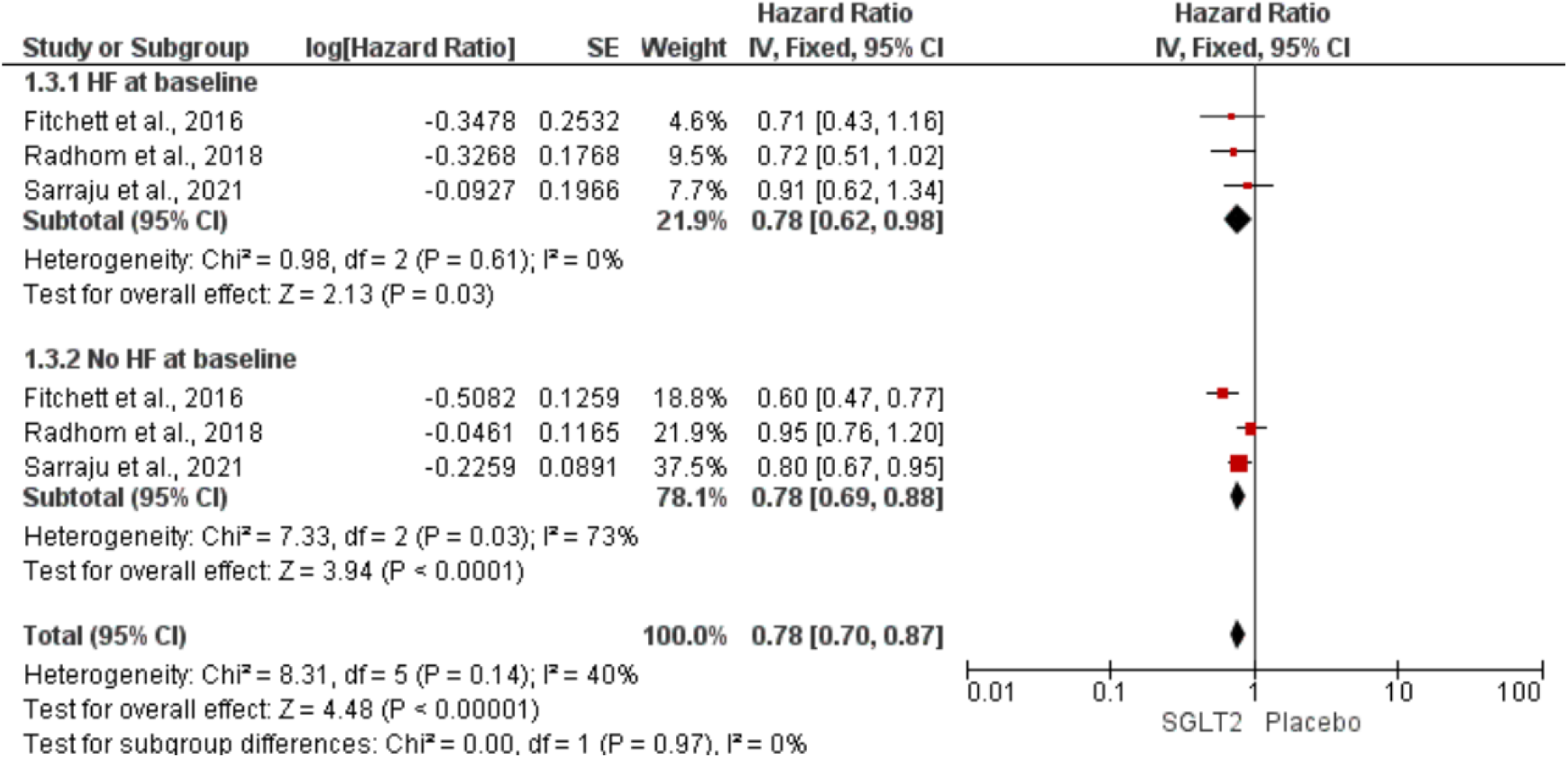
Sub-group analysis for cardiac death based on HF status at baseline.

### All-cause mortality

SGLT2 inhibitors significantly reduced all-cause mortality among participants [HR: 0.87 (0.78–0.96); P = 0.04; I^2^ = 55%] (Figure 7). The findings were consistent in patients with and without heart failure at baseline [*χ*2 = 0.08, df = 1; P = 0.78), I^2^= 0%] as demonstrated by sub-group analysis (Figure 8).

**Figure 7.**
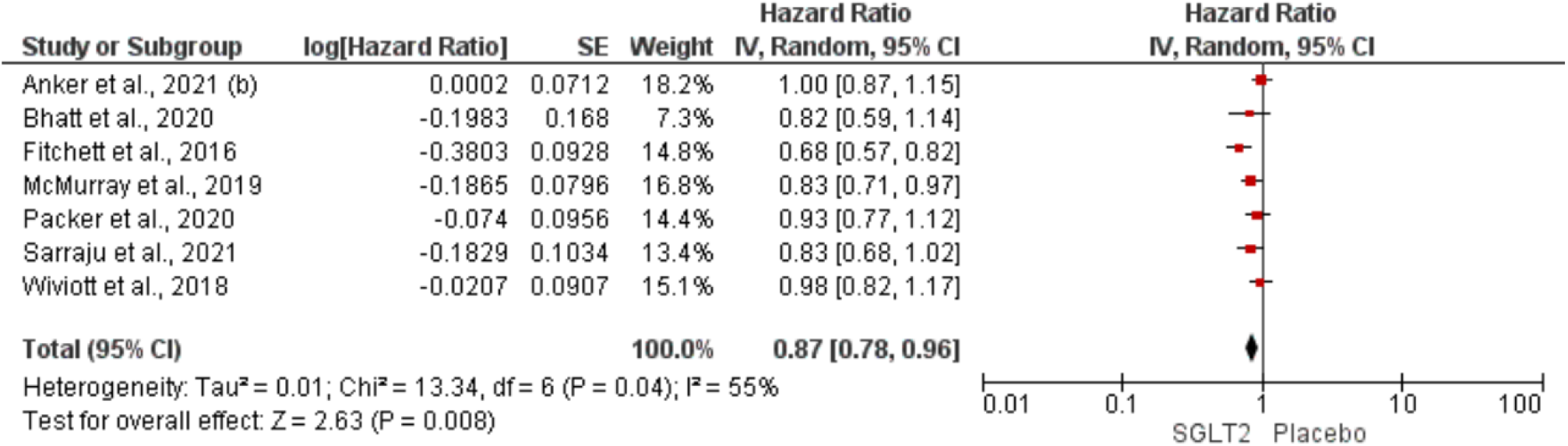
Forest plot displaying effect size for all-cause mortality among all patients.

**Figure 8.**
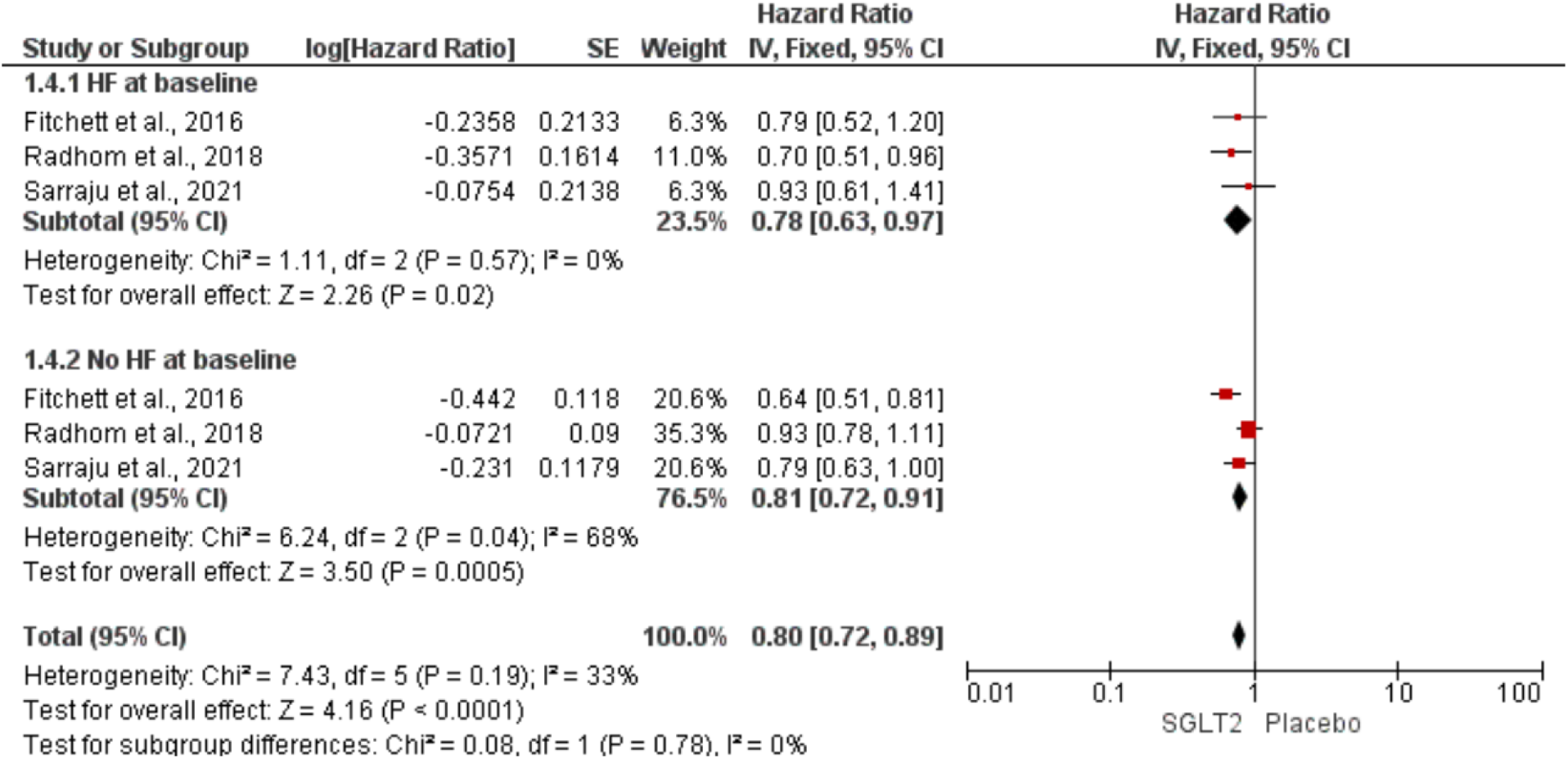
Sub-group analysis for all-cause mortality based on HF status at baseline.

### Renal composite outcome

SGLT2 inhibitor reduced the occurrence of the composite renal endpoint [HR: 0.6\70 (0.– 0.83); P < 0.001; I^2^ = 55%] (Figure 9). This finding was consistent in both post-hoc analyses for patients with and without heart failure [*χ*2 = 0.26, df = 1; P = 0.61), I^2^= 0%] (Figure 10).

**Figure 9.**
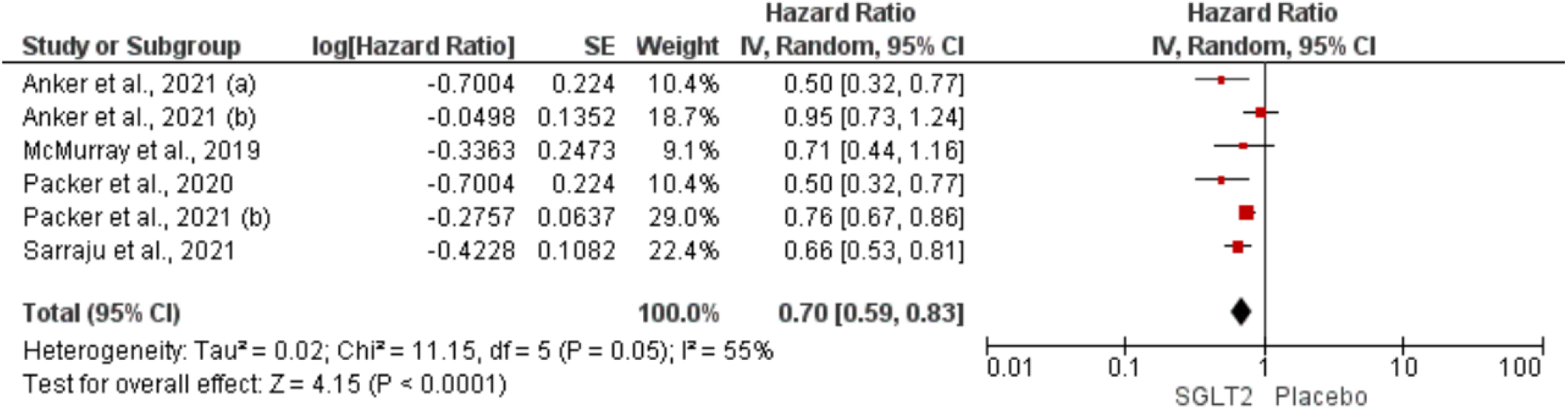
Forest plot displaying effect size for renal composite outcome among all patients.

**Figure 10.**
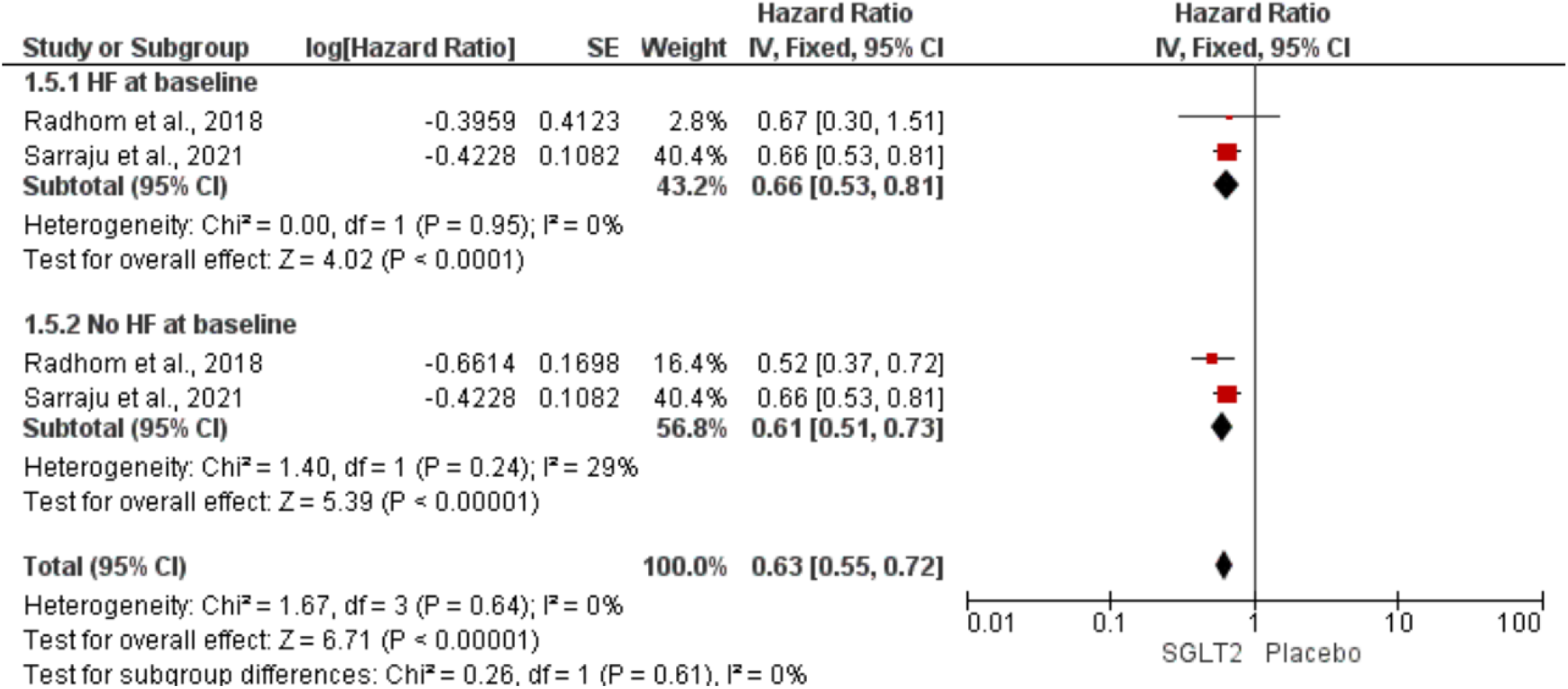
Sub-group analysis for renal composite outcome based on HF status at baseline.

### Assessing publication bias

In the funnel plot (Figure 11), the data points represent the RCTs included in the study. They all lie within the 95% CI. The data points are predominantly located in the apex of the 95% CI with some predominantly located on the left. The depicted asymmetry indicates there might have been some bias in the study that might overestimate the intervention’s effect and this should be kept in mind when used by clinicians as a reference.

**Figure 11.**
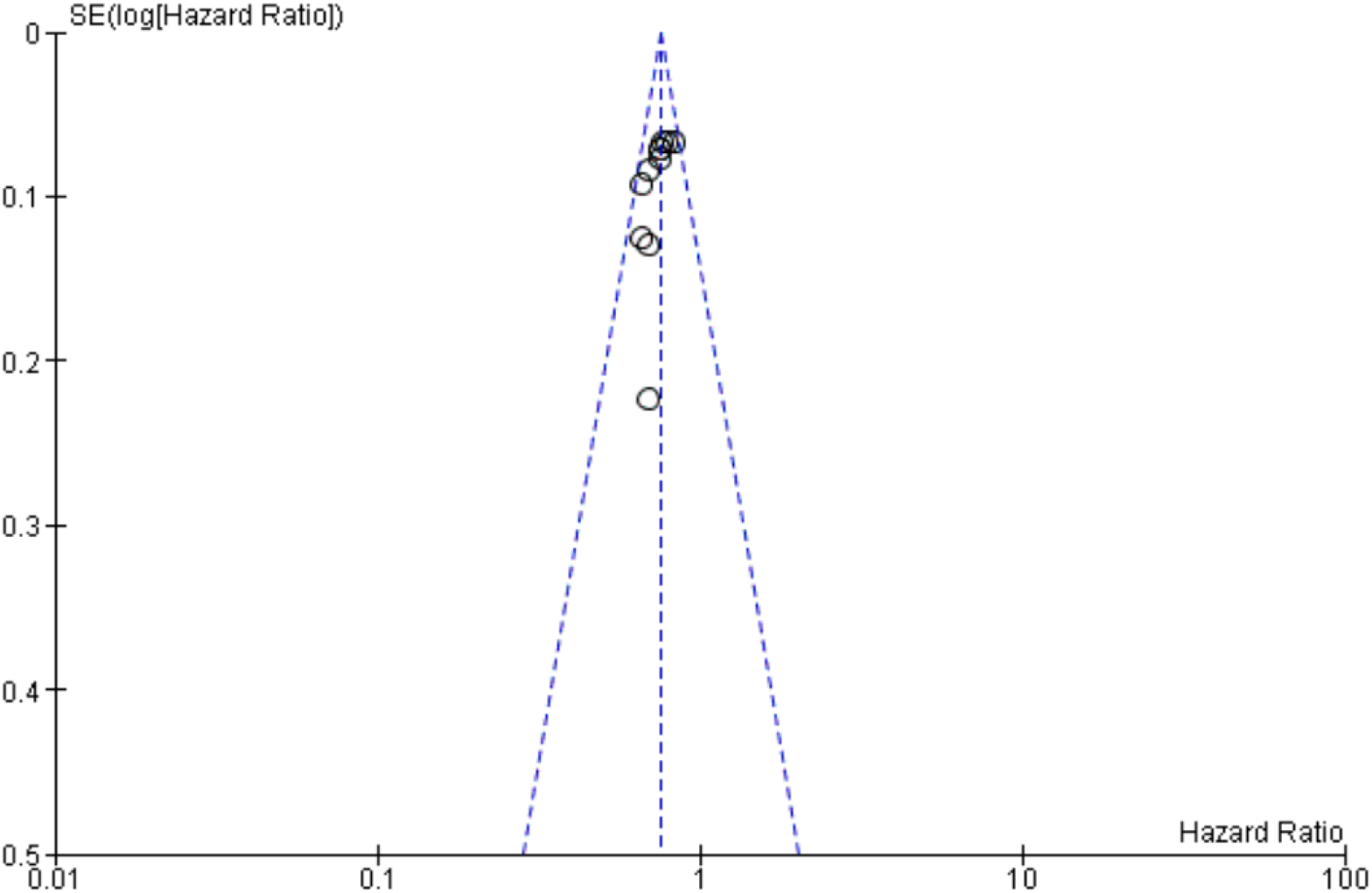
Funnel plot to assess publication bias.

## Discussion

SGLT2 inhibitors significantly decreased the risk of cardiovascular mortality, HF hospitalisation, all-cause mortality, and renal outcomes in this extensive investigation involving over 42 000 HF patients. These results were true when the HFrEF population was examined alone and when the patients were divided into groups based on their diabetes status. However, this tendency was not observed in the HFpEF subgroup. Due to the limited and inconsistent availability of data, this discovery should be viewed as exploratory rather than conclusive. SGLT2 inhibitors were not shown to increase the probability of major adverse events or discontinuation owing to adverse events compared to placebo.

All HF-specific studies revealed statistically significant decreases in the composite of cardiovascular death or total HF hospitalisation. These trials’ meta-analysis shows a substantial decrease in cardiovascular and all-cause mortality. The reduction in mortality in HF patients was further supported by findings from post-hoc analyses of cardiovascular outcome studies. We report that the reduction in mortality advantage is true independent of DM status.

The funnel plot was used to assess publication bias, where all the selected RCT lie within the 95% confidence interval but predominantly towards the left side which might indicate presence of bias to some extend. Finally, despite the fact that methodological heterogeneity was accounted for using a random-effects model, some variations between studies may make interpretation more difficult. This includes the use of various SGLT2 inhibitor subtypes, varying sex ratios, variations in background therapies, baseline risk severity of patients included, and variances in safety and renal endpoint definitions among studies.

The results of this meta-analysis demonstrate that SGLT2 inhibitors significantly lower mortality, renal outcomes, and HF hospitalisations. These results were true for individuals with HFrEF as well as for diabetics and non-diabetics. In patients with HFpEF, there was no upward trend toward improvement. Our results confirm that all HF patients, regardless of diabetes status, may benefit from SGLT2 inhibitors; however, the benefits for patients with preserved ejection fraction are still debatable; we require more RCT to study it more precisely. The results of this study can be useful for clinicians in making decision whether to incorporate SGLT2 inhibitors in their treatment plan.

## Data Availability

All data produced in the present study are available upon reasonable request to the authors.

## Declarations

### Competing interests

The authors declare that they have no competing interests.

### Funding

None.

### Authors’ contributions

SA conceptualised the meta-analysis and prepared the initial draft of the manuscript. AZ supported writing of the draft and oversaw its critical review and editing. *Both SA and AZ contributed equally to this work as co-first-authors. SK, KT, and AT undertook the literature search and helped prepare the manuscript draft. All authors read and approved the final manuscript.

## Acknowledgements

We would like to express our gratitude to Ms Qurat ul Ain for her support with the statistical analysis.

## Notes

### Competing Interest Statement

The authors have declared no competing interest.

### Funding Statement

This study did not receive any funding.

