## Supplementary Materials for "Effectiveness of SGLT2 inhibitor therapy in treatment of Heart failure: A Meta-Analysis"

**Table S1.** Search strategy for databases and their respective outputs.

Sodium Glucose Transporter 2 Inhibitors OR sodium-glucose co-transporter inhibitor OR SGLT-2 Inhibitors OR SGLT 2 Inhibitors OR SGLT2 Inhibitors OR Gliflozins OR tofogliflozin OR sotagliflozin OR empagliflozin OR canagliflozin OR dapagliflozin OR ertugliflozin OR luseugliflozin OR ipragliflozin OR remogliflozin OR sergliflozin

AND

Cardiac Failure OR Myocardial Failure OR Congestive Heart Failure OR heart failure OR OR Heart Decompensation

| PubMed | ("sodium glucose transporter 2 inhibitors"[Pharmacological Action] OR "sodium glucose transporter 2 inhibitors"[MeSH Terms] OR "sodium glucose transporter 2 inhibitors"[All Fields] OR "sodium glucose transporter 2 inhibitors"[All Fields] OR ("sodium-glucose"[All Fields] AND ("symporters"[MeSH Terms] OR "symporters"[All Fields] OR ("co"[All Fields] AND "transporter"[All Fields]) OR "co transporter"[All Fields]) AND ("antagonists and inhibitors"[MeSH Subheading] OR ("antagonists"[All Fields] AND "inhibitors"[All Fields]) OR "antagonists and inhibitors"[All Fields] OR "inhibitors"[All Fields] OR "inhibitor"[All Fields] OR "inhibitor s"[All Fields])) OR ("sodium glucose transporter 2 inhibitors"[Pharmacological Action] OR "sodium glucose transporter 2 inhibitors"[MeSH Terms] OR "sodium glucose transporter 2 inhibitors"[All Fields] OR "sglt 2 inhibitors"[All Fields]) OR ("sodium glucose transporter 2 inhibitors"[Pharmacological Action] OR "sodium glucose transporter 2 inhibitors"[MeSH Terms] OR "sodium glucose transporter 2 inhibitors"[All Fields] OR "sglt 2 inhibitors"[All Fields]) OR ("sodium glucose transporter 2 inhibitors"[Pharmacological Action] OR "sodium glucose transporter 2 inhibitors"[MeSH Terms] OR "sodium glucose transporter 2 inhibitors"[All Fields] OR ("sglt2"[All Fields] AND "inhibitors"[All Fields]) OR "sglt2 inhibitors"[All Fields]) OR ("sodium glucose transporter 2 inhibitors"[Pharmacological Action] OR "sodium glucose transporter 2 inhibitors"[MeSH Terms] OR "sodium glucose transporter 2 inhibitors"[All Fields] OR "gliflozin"[All Fields] OR "gliflozins"[All Fields]) OR  ("empagliflozin"[Supplementary Concept] OR "empagliflozin"[All Fields]) OR ("canagliflozin"[MeSH Terms] OR "canagliflozin"[All Fields]) OR ("dapagliflozin"[Supplementary Concept] OR "dapagliflozin"[All Fields] OR "dapagliflozin s"[All Fields]) OR ("ertugliflozin"[Supplementary Concept] OR "ertugliflozin"[All Fields]) OR ("ipragliflozin"[Supplementary Concept] OR "ipragliflozin"[All Fields]) OR "remogliflozin"[All Fields] OR ("sergliflozin"[Supplementary Concept] OR "sergliflozin"[All Fields])) AND ("heart failure"[MeSH Terms] OR ("heart"[All Fields] AND "failure"[All Fields]) OR "heart failure"[All Fields] OR ("cardiac"[All Fields] AND "failure"[All Fields]) OR "cardiac failure"[All Fields] OR ("heart failure"[MeSH Terms] OR ("heart"[All Fields] AND "failure"[All Fields]) OR "heart failure"[All Fields] OR ("myocardial"[All Fields] AND "failure"[All Fields]) OR "myocardial failure"[All Fields]) OR ("heart failure"[MeSH Terms] OR ("heart"[All Fields] AND "failure"[All Fields]) OR "heart failure"[All Fields] OR ("congestive"[All Fields] AND "heart"[All Fields] AND "failure"[All Fields]) OR "congestive heart failure"[All Fields]) OR ("heart failure"[MeSH Terms] OR ("heart"[All Fields] AND "failure"[All Fields]) OR "heart failure"[All Fields]) OR ("heart failure"[MeSH Terms] OR ("heart"[All Fields] AND "failure"[All Fields]) OR "heart failure"[All Fields] OR ("heart"[All Fields] AND "decompensation"[All Fields]) OR "heart decompensation"[All Fields])) | 2398 |
| --- | --- | --- |
| PsycInfo | **Any Field**: Sodium Glucose Transporter 2 Inhibitors *OR* **Any Field**: sodium-glucose co-transporter inhibitor *OR* **Any Field**: SGLT-2 Inhibitors *OR* **Any Field**: SGLT 2 Inhibitors *OR* **Any Field**: SGLT2 Inhibitors *OR* **Any Field**: Gliflozins *OR* **Any Field**: tofogliflozin *OR* **Any Field**: sotagliflozin *OR* **Any Field**: empagliflozin *OR* **Any Field**: canagliflozin *OR* **Any Field**: dapagliflozin *OR* **Any Field**: ertugliflozin *OR* **Any Field**: luseugliflozin *OR* **Any Field**: ipragliflozin *OR* **Any Field**: remogliflozin *OR* **Any Field**: sergliflozin *AND* **Any Field**: Cardiac Failure *OR* **Any Field**: Myocardial Failure *OR* **Any Field**: Congestive Heart Failure *OR* **Any Field**: heart failure *OR* **Any Field**: OR Heart Decompensation | 110 |
| Google Scholar | "Sodium Glucose Transporter 2 Inhibitors" AND "cardiac failure" | 55 |

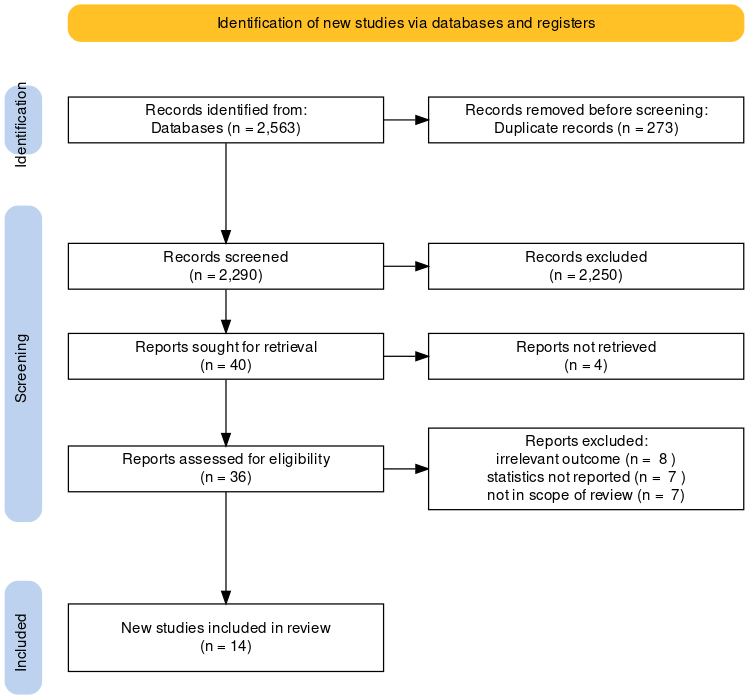

**Figure S1.** PRISMA flowchart outlining the paper screening process.
